# Sequential viral introductions and spread of BA.1 drove the Omicron wave across Pakistani provinces

**DOI:** 10.1101/2023.03.25.23287718

**Authors:** Ali Raza Bukhari, Javaria Ashraf, Akbar Kanji, Yusra Abdul Rahman, Nídia S. Trovão, Peter M. Thielen, Maliha Yameen, Samiah Kanwar, Waqasuddin Khan, Furqan Kabir, M. Imran Nisar, Brian Merritt, Rumina Hasan, David Spiro, Zeba Rasmussen, Uzma Bashir Aamir, Zahra Hasan

**Affiliations:** Department of Pathology and Laboratory Medicine, Aga Khan University, Karachi-74800, Pakistan; Johns Hopkins University Applied Physics Laboratory, 11100 Johns Hopkins Road, Laurel, MD 20723, USA; Fogarty International Center, U.S. National Institutes of Health, 16 Center Drive, Bethesda, MD 20892, USA; Department of Pediatrics and Child Health, Medical College, Aga Khan University, Karachi-74800, Pakistan; World Health Organization country office, Park Road, Chak Shahzad, Islamabad, Pakistan

**Keywords:** Omicron, variants of concern, SARS-CoV-2, phylogenetics, transmission dynamics

## Abstract

**Background:** COVID-19 waves caused by specific SARS-CoV-2 variants have occurred globally at different times. We focused on Omicron variants to understand the genomic diversity and phylogenetic relatedness of SARS-CoV-2 strains in various regions of Pakistan.

**Methods:** We studied 276,525 COVID-19 cases and 1,041 genomes sequenced from December 2021 to August 2022. Sequences were analyzed and visualized using phylogenetic trees.

**Results:** The highest case numbers and deaths were recorded in Sindh and Punjab, the most populous provinces in Pakistan. Omicron variants comprised 95% of all genomes, with BA.2 (34.2%) and BA.5 (44.6%) predominating. The first Omicron wave was associated with the sequential identification of BA.1 in Sindh, then Islamabad Capital Territory, Punjab, Khyber Pakhtunkhwa (KP), Azad Jammu Kashmir (AJK), Gilgit-Baltistan (GB) and Balochistan. Phylogenetic analysis revealed Sindh to be the source of BA.1 and BA.2 introductions into Punjab and Balochistan during early 2022. BA.4 was first introduced in AJK and BA.5 in Punjab. Most recent common ancestor (MRCA) analysis revealed relatedness between the earliest BA.1 genome from Sindh with Balochistan, AJK, Punjab and ICT, and that of first BA.1 from Punjab with strains from KPK and GB.

**Conclusions:** Phylogenetic analysis provides insights into the introduction and transmission dynamics of the Omicron variant in Pakistan, identifying Sindh as a hotspot for viral dissemination. Such data linked with public health efforts can help limit surges of new infections.

## Introduction

The rise and fall of coronavirus disease 2019 (COVID-19) cases since the emergence of severe acute respiratory syndrome coronavirus 2 (SARS-CoV-2) in December 2019 has driven the need for monitoring of variants through genomic surveillance [1]. The global burden of COVID-19 is not fully known although greater than 754 million cases have been reported as of February 3, 2022. Pakistan has reported about 1.5 million cases of COVID-19 and nearly 31,000 deaths [2, 3]. COVID-19 vaccines have had a significant impact on controlling both morbidity and mortality from COVID-19 [4, 5]. The proportion of deaths to cases changed greatly throughout the pandemic with reduced disease severity after vaccines were introduced and the evolution of SARS-CoV-2 variants of concern (VoC) toward higher transmissibility and lower pathogenicity. The epidemiology of COVID-19 has been informed by the reported number of cases, deaths, hospitalizations, and viral genomic sequencing. This information has varied greatly between high- and low-income countries of similar population sizes; explanatory factors include lack of resources needed for gathering metadata, PCR testing and genomic sequencing [6].

SARS-CoV-2 VoCs have shown a trend toward increased transmissibility, leading to strain-specific increases in COVID-19 cases [7]. The first VoC was the Alpha variant or B.1.1.7, followed by Beta/B.1.351 and Delta/B.1.617.2 in 2021. VoCs which emerged after the introduction of vaccinations were more effective at evading host immunity driven by both natural infection and vaccinations [8]. This was evidenced by Omicron and its subvariants coming to dominate over other lineages across the globe by the end of 2021 [9, 10]. Omicron has distinct subvariants, five of which include BA.1, BA.2, BA.3, BA.4 and BA.5 [11]. BA.1 was first identified in South Africa and Botswana on November 26, 2021 [12]. By January 2022, BA.1 made up 81% of cases in South Africa, rapidly decreasing to 45% in February 2022. BA.5 was first identified in South Africa on January 21, 2022, and spread elsewhere, particularly in Europe, by April 2022. Soon after this time, BA.4 and BA.5 variants became predominant [13].

Understanding the relationship between COVID-19 rates and SARS-CoV-2 variants at the regional level is important for management of the public health response. The identification of new SARS-CoV-2 variants and their prevalence in different regions can help make public health strategies, such as vaccination campaigns, testing protocols, and contact tracing efforts. By monitoring the circulation of SARS-CoV-2 variants and their impact on COVID-19 rates, public health officials can make decisions about the allocation of resources and implementation of preventive measures to limit the spread of the virus.

Pakistan has a population of approximately 220 million people. It consists of seven distinct regional territories: Azad Jammu and Kashmir (AJK), Balochistan, Gilgit-Baltistan (GB), Khyber Pakhtunkhwa (KP), Punjab, Sindh and the Islamabad Capital Territory (ICT). The first COVID-19 case from Pakistan was reported in Sindh on February 26, 2020. The timeline of pandemic waves experienced in the country was; March to July 2020 where the G Nextstrain clade of SARS-CoV-2 dominated, October 2020 to January 2021 dominated by GR/GH clades, April to May 2021 where Alpha dominated [14], July to September 2021 where Delta was the predominant strain, and, December 2021 to February 2022 dominated by Omicron.

Here, we studied COVID-19 waves in association with cases and mortality and Omicron subvariants identified in regions across Pakistan. We also investigated the introduction and relatedness of variants in each region to understand the patterns of transmission.

## Materials and Methods

This study was approved by the Ethical Review Committee, The Aga Khan University (AKU).

### Selection of SARS-CoV-2 genomes

This was an analysis of SARS-CoV-2 genomes submitted of Pakistani origin. Sequences with collection dates between December 1, 2021 and August 14, 2022 were obtained from GISAID on September 15, 2022 “complete” and “low coverage excluded” filtering criteria. We downloaded 1041 sequences with metadata; of these, 984 were found to belong to the Omicron VoC, of which 957 had complete metadata and were used for phylogenetic analysis.

### COVID-19 cases and mortality data

COVID-19 case data for the period December 1, 2021 to August 14, 2022 were downloaded from the John Hopkins coronavirus resource center [3] and used for analysis. COVID-19 mortality data were downloaded from Pakistan’s official COVID-19 page (https://covid.gov.pk/) accessed on September 1, 2022.

### Phylogenetic tree and analysis

FASTA files of the 984 Omicron sequences with corresponding metadata (age, gender, date of collection and location) were used for phylogenetic tree reconstruction using the augur pipeline [15]. Out of the total (984), 957 genomic sequences qualified for phylodynamic mapping.

Full length SARS-CoV-2 genomes of Omicron subvariants were aligned using the MAFFT alignment tool [16]. Multiple Sequence Alignment (MSA) files generated from MAFTT were used for a maximum likelihood (ML) phylogenetic tree through IQ-TREE2 [17]. By applying a generalized midpoint rooting strategy, rooting of the tree was carried out with branch length variance using TreeTime [18]. The tree was visualized and edited in Figtree *v*. 1.4.4 (http://tree.bio.ed.ac.uk). The final tree with annotated nodes and metadata was exported to the phylodynamic visualizing tool Auspice [19].

### Statistical Analysis

Demographic results are presented in mean ± SD. Kruskal–Wallis statistical tests were used to analyze statistical significance, with p-value less than 0.05 considered statistically significant. Graph Pad Prism *v*. 5.0 (http://www.graphpad.com) was used for statistical analysis.

## Results

### Infections and deaths in different regions of Pakistan during the fifth COVID-19 wave

A total of 276,525 cases were reported between December 2021 and August 2022 [20]. The region-wide distribution of COVID-19 cases is depicted in Figure 1, which also depicts the population density of each of the 7 regions studied. Sindh reported the most cases (41.7%), followed by Punjab with 27.1%, 15.2% from KP, 11% from ICT, 3.4% from AJK, 0.9% from Balochistan and 0.6% from GB of total cases.

**Figure 1.**
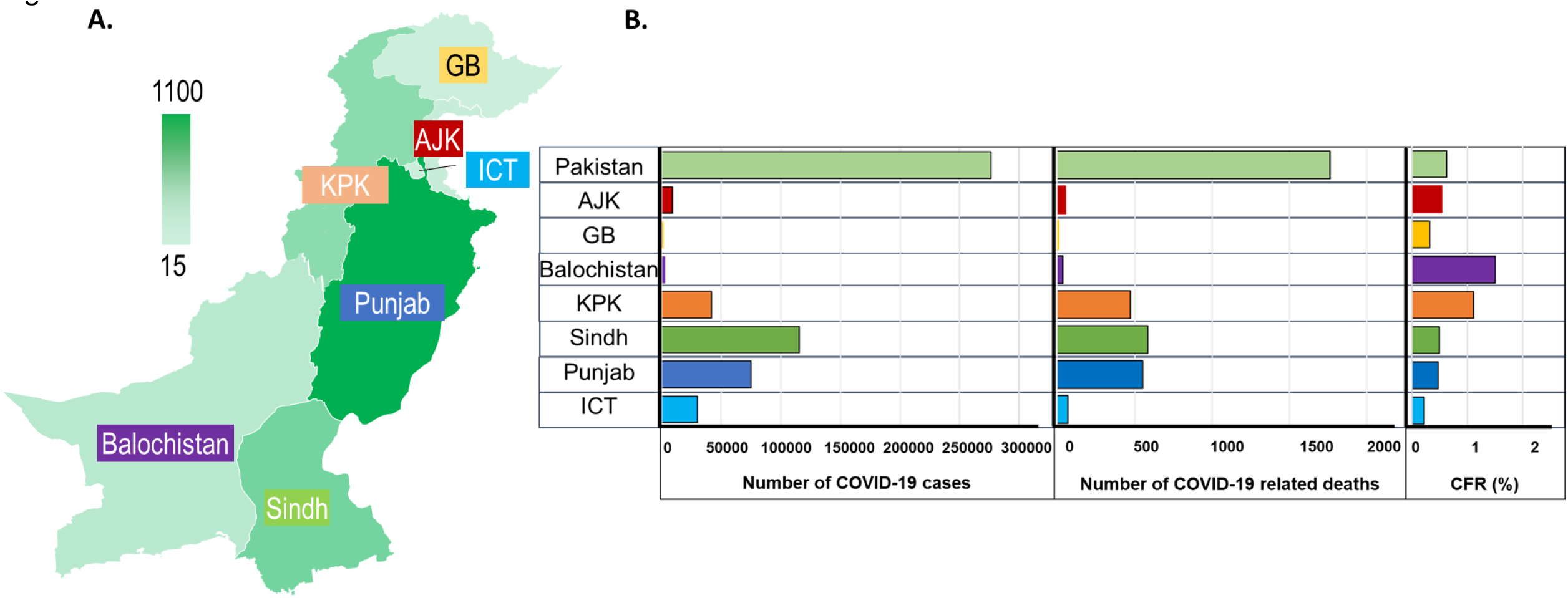
Population density, COVID-19 cases, deaths and CFR% across Pakistan. The graph depicts nationwide data from December 1, 2021, until August 14, 2022. A. The region-wise population density of Azad Jammu and Kashmir (AJK), Balochistan, Gilgit-Baltistan (GB), Islamabad Capital Territory (ICT), Khyber Pakhtunkhwa (KPK), Punjab and Sindh is presented. The scale bar displays population values in millions of persons shaded by color. B. Left panel x-axis shows number of COVID-19 cases, middle panel shows number of COVID-19 related deaths and right panel shows CFR% of regions represented by color: ICT (light blue), Punjab (dark blue), Sindh (green), KPK (orange), Balochistan (purple), GB (yellow) and AJK (red).

A total of 1,766 COVID-19 deaths were reported during the study period (Figure 1). Sindh reported the greatest (33.1% of total deaths) followed by Punjab (31.1%), KP (26.7%), ICT (3.9%), AJK (2.9%), Balochistan (2.0%) and GB (0.3%). The overall case fatality ratio percentage (CFR%) over this period was 0.6%. It was highest in Balochistan (1.5%), followed by KP (1.1%), AJK (0.5%), Sindh (0.5%), Punjab (0.5%), GB (0.3%) and ICT (0.2%).

COVID-19 case numbers rose rapidly from under 1,000 to more than 10,000 per week across Pakistani regions between the end of January and beginning of February 2022, with 22,000 cases being reported in the last week of January 2022 in Sindh alone (Figure 2). COVID-19 peaks occurred sequentially in other provinces; ICT by the last week of January 2022; Punjab, KP, Balochistan and AJK by the first week of February 2022; and GB by the second week of February 2022. Cases declined around early March 2022, reaching a few hundred cases per week by June 2022. Another slight rise in cases was observed in Sindh (early July 2022) and Punjab (early August 2022). A similar trend was also observed in other regions. Of note, a rise in COVID-19 cases was observed in Sindh ahead of the other regions of Pakistan.

**Figure 2.**
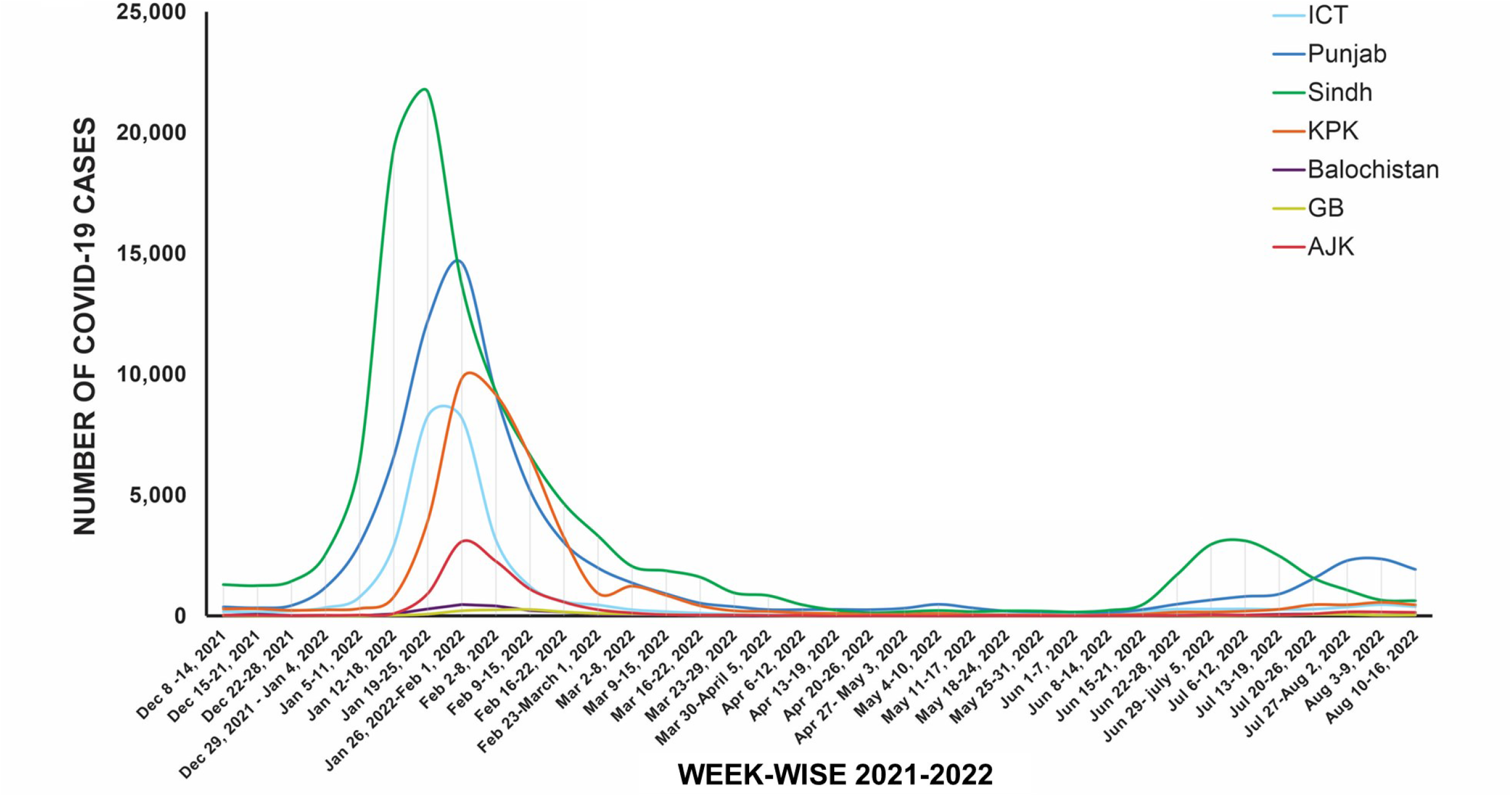
Trend of COVID-19 cases in Pakistan. The figure depicts the weekly count of COVID-19 cases through the period from December 1, 2021, until August 14, 2022. Cases are represented by region ICT (light blue), Punjab (dark blue), Sindh (green), Khyber Pakhtunkhwa (KPK, orange), Balochistan (purple), GB (yellow) and AJK (red).

The mean age of COVID-19 cases across Pakistan was 39 SD19 years. For each region the mean age was: ICT 38 SD19, Punjab 39 SD21, Sindh 40 SD19, KP 32 SD19, Balochistan 48 SD17, AJK 38 SD19 and GB 43 SD24 years. There was no significant difference between age groups of COVID-19 cases across the regions.

### Heterogeneity in data submission across regions of Pakistan

We investigated the association of COVID-19 waves with SARS-CoV-2 Omicron variants. We performed phylogenetic analysis of the 957 Omicron genomes available in relation to their date and location of sample collection. The monthly rate of submissions across the study period was not uniform (Supplementary Figure 1 and 2). More genomes were submitted in March, June, July and August 2022. There was variability in the location of the submissions. Provinces with laboratories with genomic surveillance capacity had greater representation; Sindh (n=364), ICT (376) and Punjab (n=117) contributed more than 80% of total submissions, with limited representation from GB (n=43), KP (n=61), and AJK (n=63). The fewest SARS-CoV-2 genomes were from Balochistan (n=14).

The pango lineage distribution was A (0.3%), AY (4.7%), B (1.7%), B.1 (0.8%), BA.1 (Nextstrain 21K; 16.6%), BA.2 (Nextstrain clade 21L; 32.2%), BA.4 (Nextstrain 22A; 3.6%) and BA.5 (Nextstrain 22B; 40.3%).

We also investigated the association between variants and age of COVID-19 cases; we divided the data for sequences available into 4 groups (≤ 18 years, 19-40 years, 41-55 years and ≥ 56 years). For each of the SARS-CoV-2 variants, we found the greatest number of cases to be in those aged 19-40 years, p<0.0001 (Table 1).

**Table 1.** Distribution of SARS-CoV2 lineages according to different age groups. The age-group wise SARS-CoV2 variants frequency is presented. The Kruskal-Wallis test was performed to study association between the age groups for every variant and p-value <0.05 is considered significant.

| **SARS-CoV2 variant**<br>**(number, %)** | **Age**<br>**(years)** | **number of**<br>**variants in each**<br>**group** | **% of variant in each**<br>**group (%)** | **p-value (Kruskal-**<br>**Wallis)** |
| --- | --- | --- | --- | --- |
| **A**<br>**(n=3; 0.3%)** | ≤18 | 0 | 0.0 | Not possible |
|  | 19-40 | 2 | 66.6 |  |
|  | 41-55 | 1 | 33.3 |  |
|  | ≥ 56 | 0 | 0.0 |  |
| **AY**<br>**(n=46, 4.7%)** | ≤18 | 3 | 6.5 | <0.0001 |
|  | 19-40 | 29 | 63.0 |  |
|  | 41-55 | 8 | 17.4 |  |
|  | ≥ 56 | 6 | 13.0 |  |
| **B**<br>**(n=17, 1.7%)** | ≤18 | 7 | 41.2 | 0.0029 |
|  | 19-40 | 7 | 41.2 |  |
|  | 41-55 | 2 | 11.8 |  |
|  | ≥ 56 | 1 | 5.9 |  |
| **B.1**<br>**(n=8, 0.8%)** | ≤18 | 3 | 37.5 | 0.0933 |
|  | 19-40 | 3 | 37.5 |  |
|  | 41-55 | 1 | 12.5 |  |
|  | ≥ 56 | 1 | 12.5 |  |
| **BA.1**<br>**(n=161, 16.6%)** | ≤18 | 12 | 7.5 | <0.0001 |
|  | 19-40 | 71 | 44.1 |  |
|  | 41-55 | 31 | 19.3 |  |
|  | ≥ 56 | 47 | 29.2 |  |
| **BA.2**<br>**(n=312, 32.2%)** | ≤18 | 34 | 10.9 | <0.0001 |
|  | 19-40 | 153 | 49.0 |  |
|  | 41-55 | 58 | 18.6 |  |
|  | ≥ 56 | 67 | 21.5 |  |
| **BA.4**<br>**(n=35, 3.6%)** | ≤18 | 5 | 14.3 | <0.0001 |
|  | 19-40 | 16 | 45.7 |  |
|  | 41-55 | 8 | 22.9 |  |
|  | ≥ 56 | 6 | 17.1 |  |
| **BA.5**<br>**(n=391, 40.3%)** | ≤18 | 40 | 10.2 | <0.0001 |
|  | 19-40 | 199 | 50.9 |  |
|  | 41-55 | 68 | 17.4 |  |
|  | ≥ 56 | 84 | 21.5 |  |

### Omicron and subvariants across regions of Pakistan

Phylogenetic analysis of Omicron subvariant sequences is depicted in Figure 3. ICT uploaded 363 sequences to GISAID, the greatest of which were BA.5 (n=161, 44.4%) and BA.2 (n=153, 42.1%). There were 139 sequences from Punjab, predominantly BA.5 (n=58, 41.7%) and BA.2 (n=52, 37.4%). The distribution of 282 genomes from Sindh included BA.5 (n=131, 46.5%), BA.2 (n=63, 22%) and BA.1 (n=76, 27%). The fewest genomes were submitted from AJK (n=63, 6%), KP (n=61, 6%) and GB (n=43, 4%); of those available, BA.2 and BA.5 were the predominant subvariants. The fewest sequences were from Balochistan (n=14, 2%), which did not report any BA.5 subvariant.

**Figure 3.**
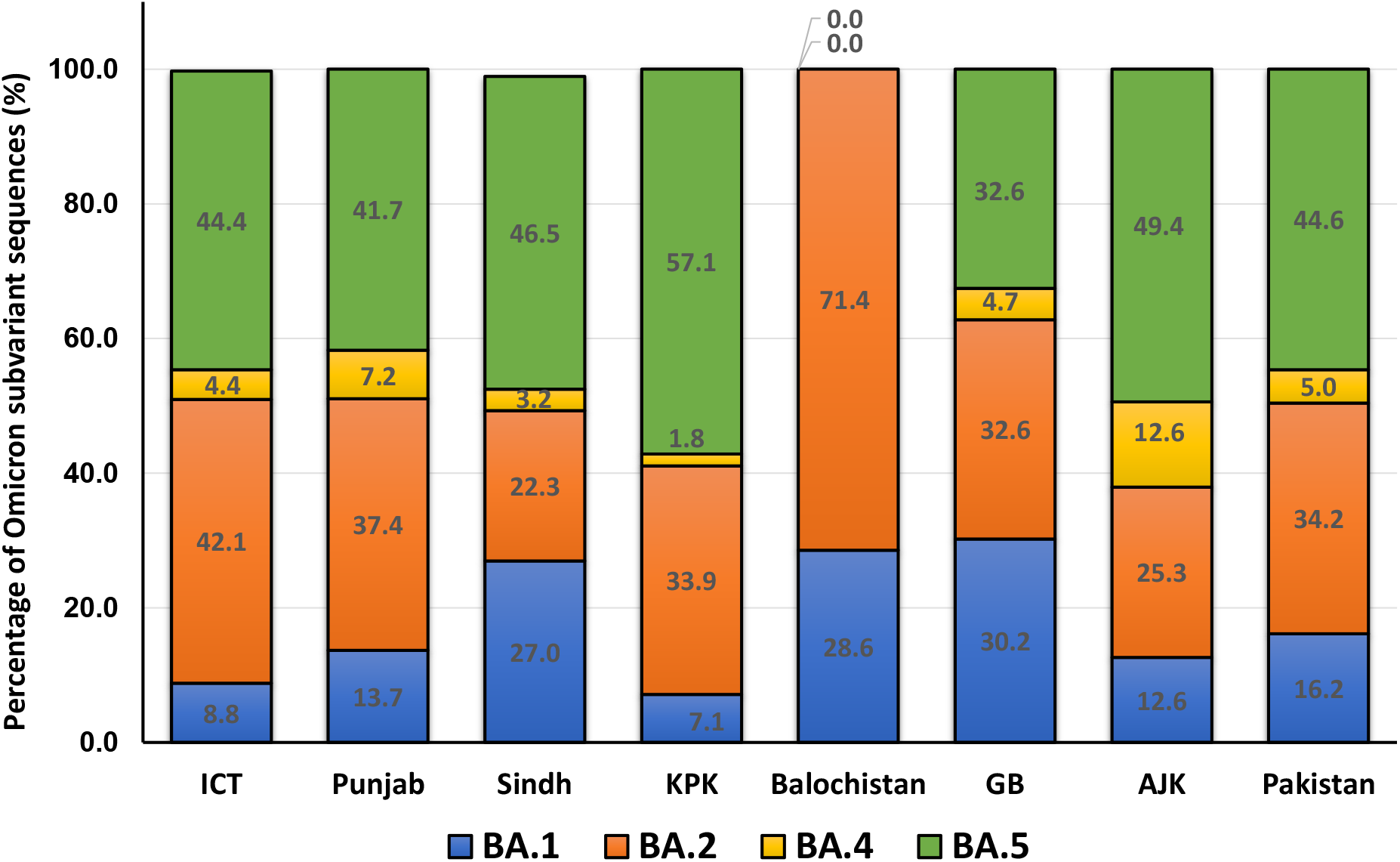
Frequency of Omicron variants across Pakistan. The graph shows all Omicron (n=984) genomes submitted from each region between December 1, 2021, and August 14, 2022. BA.1 (blue), BA.2 (orange), BA.4 (yellow) and BA.5 (green).

### Phylogenetic Analysis of SARS-CoV-2 Omicron variants from Pakistan

The phylogenetic analysis of the Omicron genomes across Pakistan is depicted in Figure 4. The phylogram presents the evolution and spread over time of BA.1, BA.2, BA.4 and BA.5 variants across the country, including sequences from ICT, Sindh, Punjab, AJK, KP, GB and Balochistan. Some were from travelers who entered Sindh from the Kingdom of Saudi Arabia (KSA, n=3), the United Arab Emirates (UAE, n =2), India (n=1), Turkey (n=1) and the United States of America (USA, n=1).

**Figure 4.**
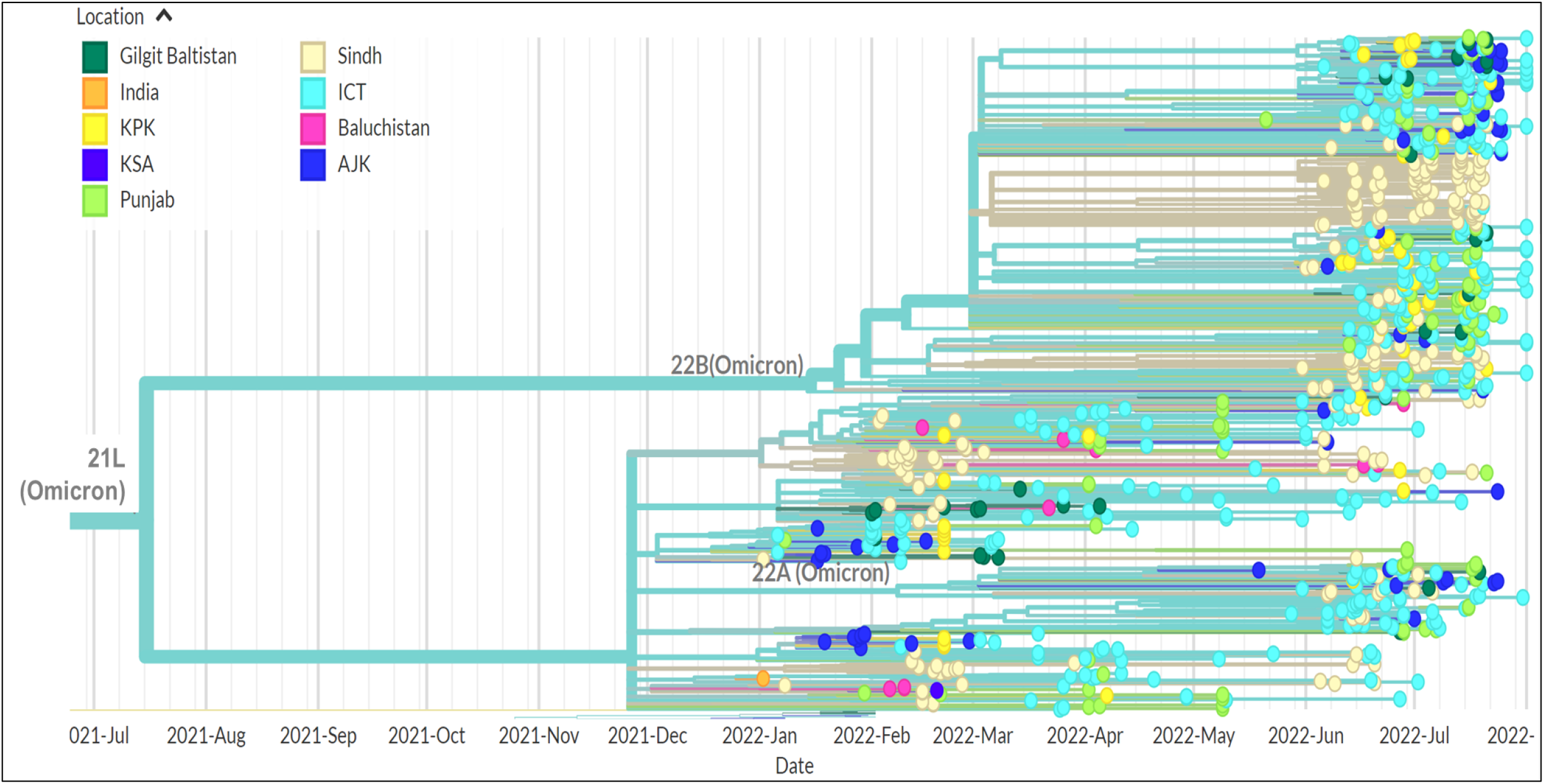
Phylogenetics of early Omicron variants in Pakistan. The tree illustrates the relatedness of 957 omicron sequences submitted from Pakistan between December 1, 2021, and August 14, 2022. The tree uses color coding to identify the travel origin of each case reported.

### Investigating the phylogeny of BA.1, BA.2, BA.4 and BA. 5 subvariants

The first BA.1 subvariant was detected on December 13, 2021, and was followed by a surge of cases. Subsequent surges were associated with the BA.2 subvariant and then the BA.4 and BA.5 subvariants. To understand this trend, we separately looked at the phylogenetics of each variant.

The first case of BA.1 was identified in Sindh (Figure 5A). The BA.1 lineage encompassed sub-lineages BA.1.1, BA.1.1.1, BA.1.1.13, BA.1.1.14, BA.1.1.18, BA.1.15, BA.1.15.1, BA.1.17, and BA.1.18. Subsequently, BA.1 was reported in Punjab, ICT, and AJK. Later, reports of BA.1 were obtained from KP (January 2022) and Balochistan (February 2022).

**Figure 5.**
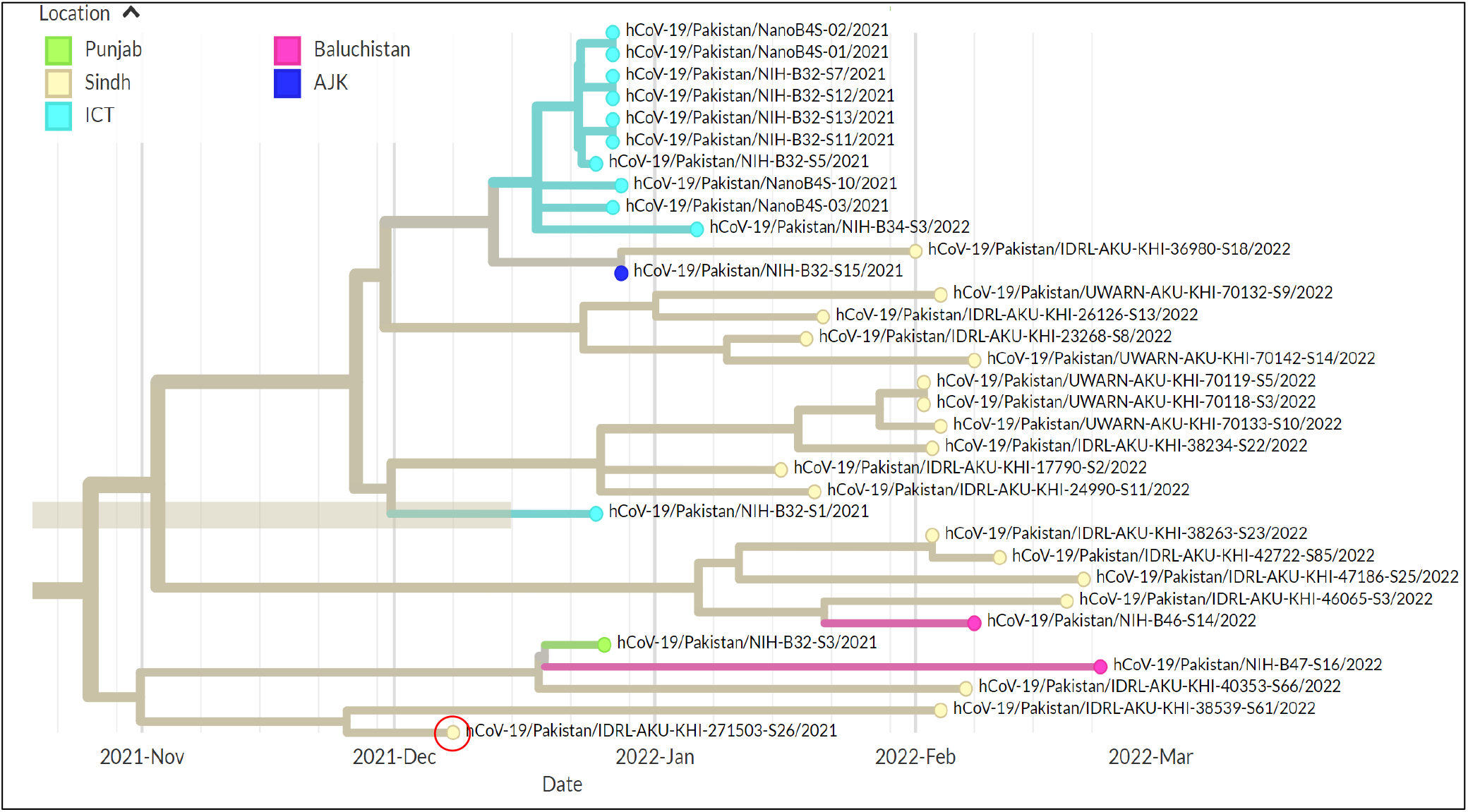
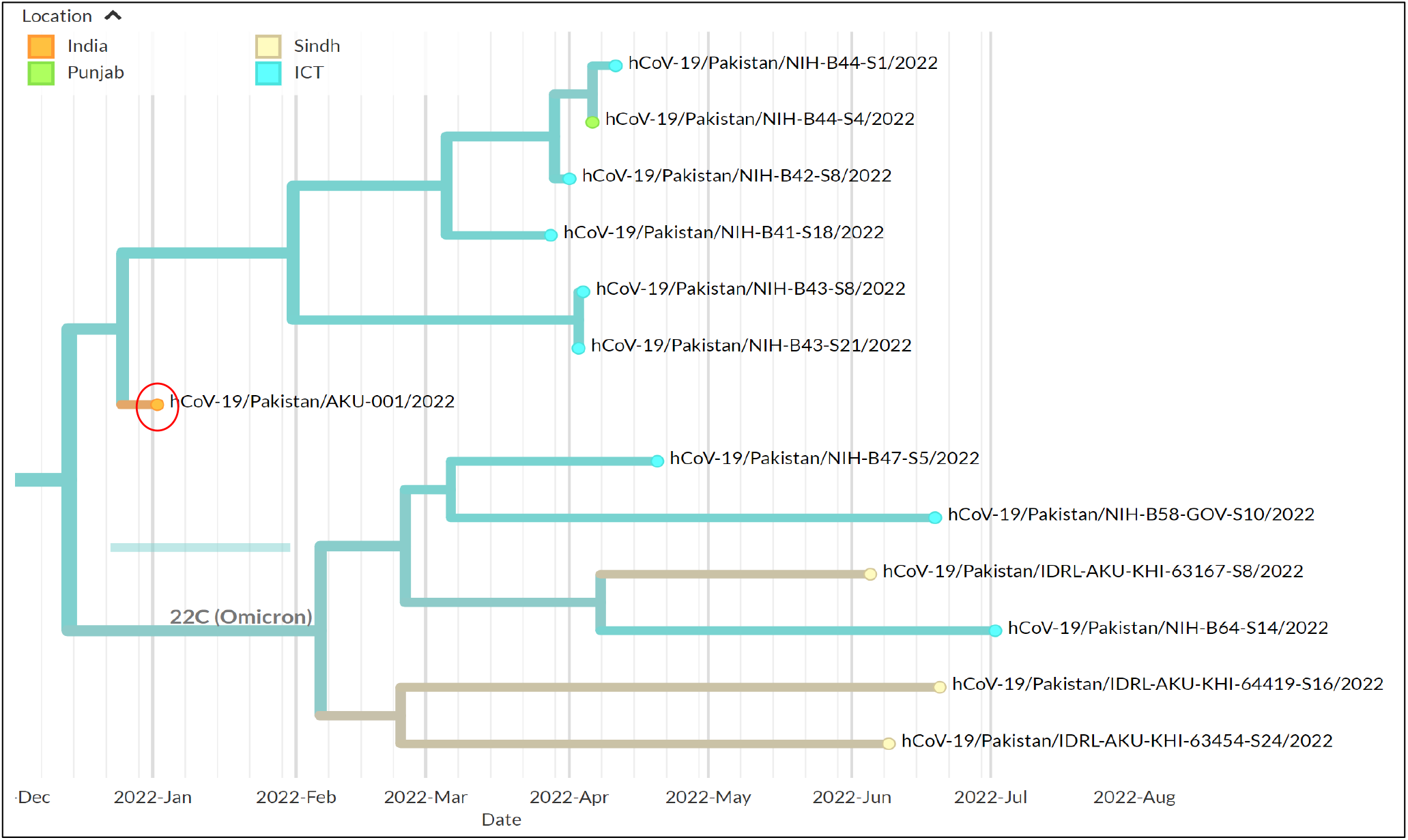
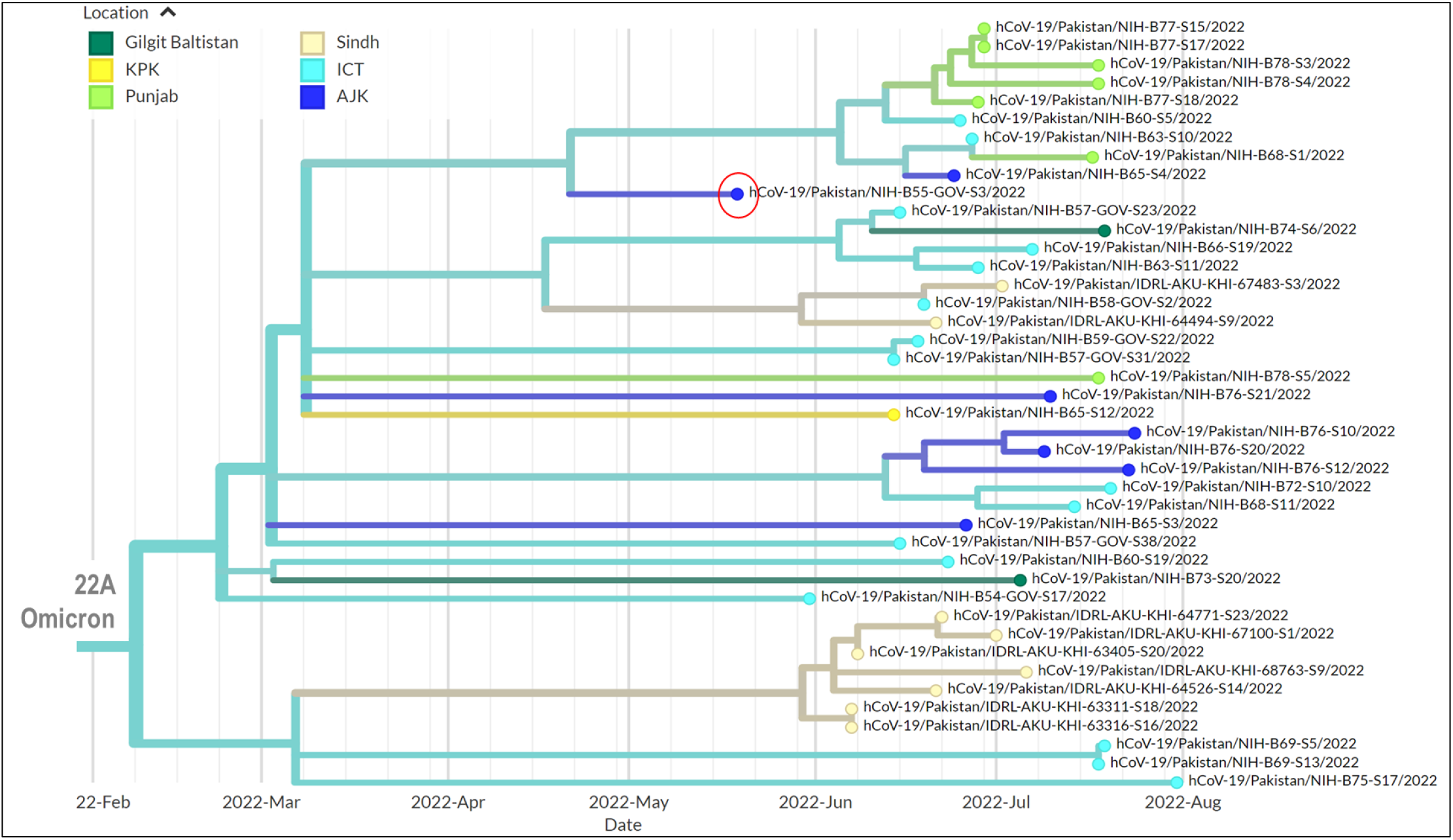
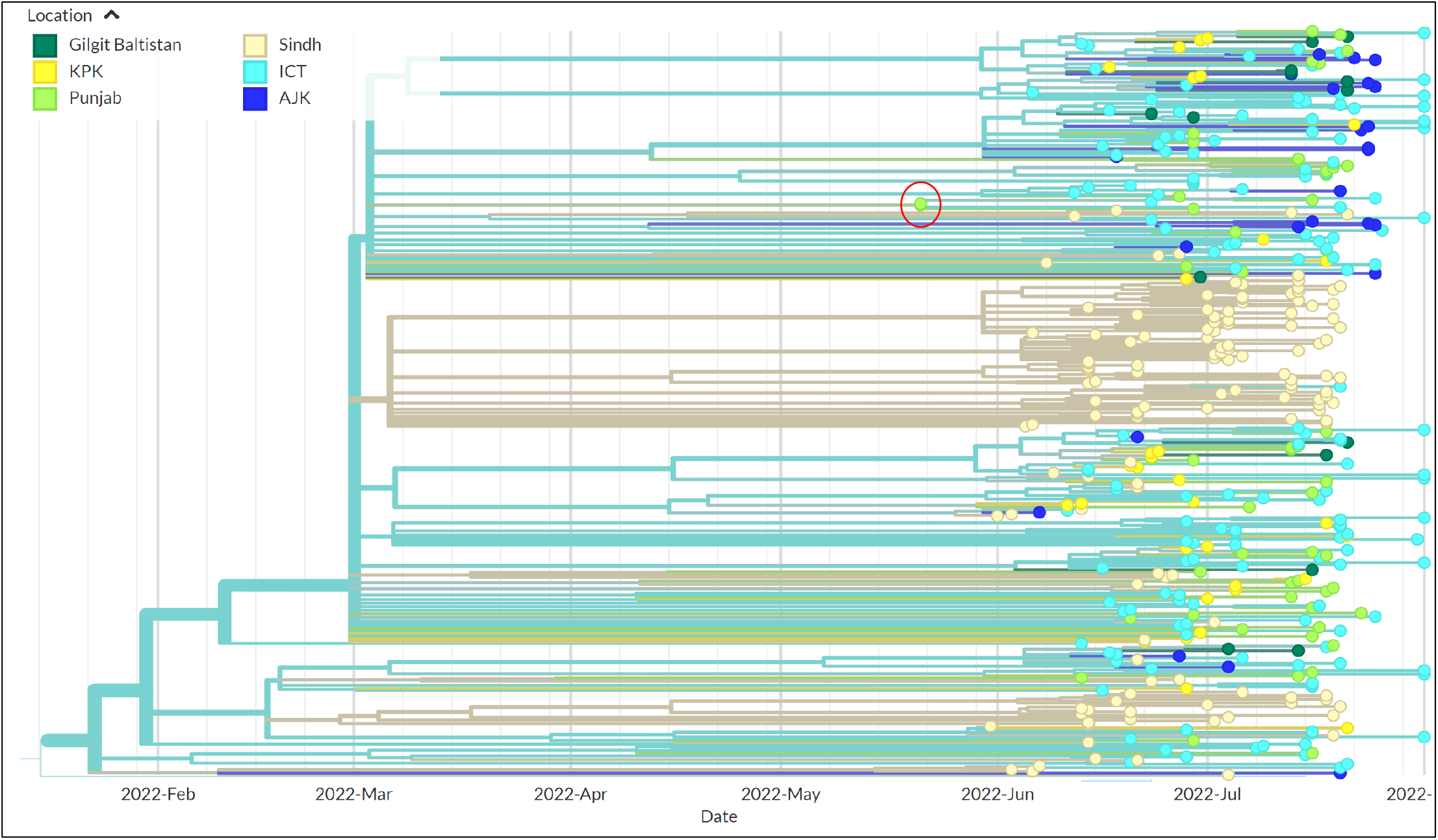
Introduction and linkage of omicron variants in Pakistan. Phylogenetic trees depict the first case report for each variant as a red circle in the identified timeline with relatedness to later isolates. Panels A-D depict the first case of each variant identified by a red circle for A, BA.1; B, BA.2; C, BA.4 and D, BA.5. Regional locations are identified by colors; Sindh in light yellow, Balochistan in pink, AJK in blue, Punjab in light green, GB in dark green and ICT, light blue. Data are presented as auspice output of the tree generated using IQ-TREE *v*. 2.2.0.

We next examined the phylogeny of the 333 BA.2 sequences found between January and July 2022. The first BA.2 subvariant was reported in January 2022 from Sindh, and clustered closely with an isolate identified in a traveler from India (Figure 5B). Subsequently, BA.2 was reported in ICT and Punjab in April 2022.

The BA.4 subvariant was first reported in AJK in May 2022, followed by reports in June 2022 onwards from ICT, Sindh, KP, Punjab and GB (Figure 5C).

The BA.5 subvariant was first reported in Punjab in May 2022, followed by reports from Sindh, ICT, AJK, KP and Gilgit (Figure 5D).

### Phylogenetic relatedness of cases reported across different regions of Pakistan

To further understand the genetic relatedness of Omicron subvariants across Pakistan, we analyzed the relatedness of sequenced genomes through the inference of the most recent common ancestor (MRCA) across all 957 genomes (Figure 6A), focusing on the first BA.1 case reported in each region. The first BA.1 case from Balochistan, with a sampling date of January 21, 2022, shared a common ancestral node with one reported earlier from Sindh on January 10, 2022 (Figure 6B). The first case from Sindh, reported on December 8, 2022, had no known association with an overseas traveler [21] (Figure 6C). The first BA.1 case from AJK (January 8, 2022) shared an ancestral node with a case from Sindh and Punjab (Figure 6D). The earliest case from GB and KP had the same MRCA and shared an ancestral node with isolates from the Punjab (Figure 6E). Similarly, the earliest BA.1 cases from ICT and Punjab shared an ancestral node with Sindh (Figure 6F).

**Figure 6.**
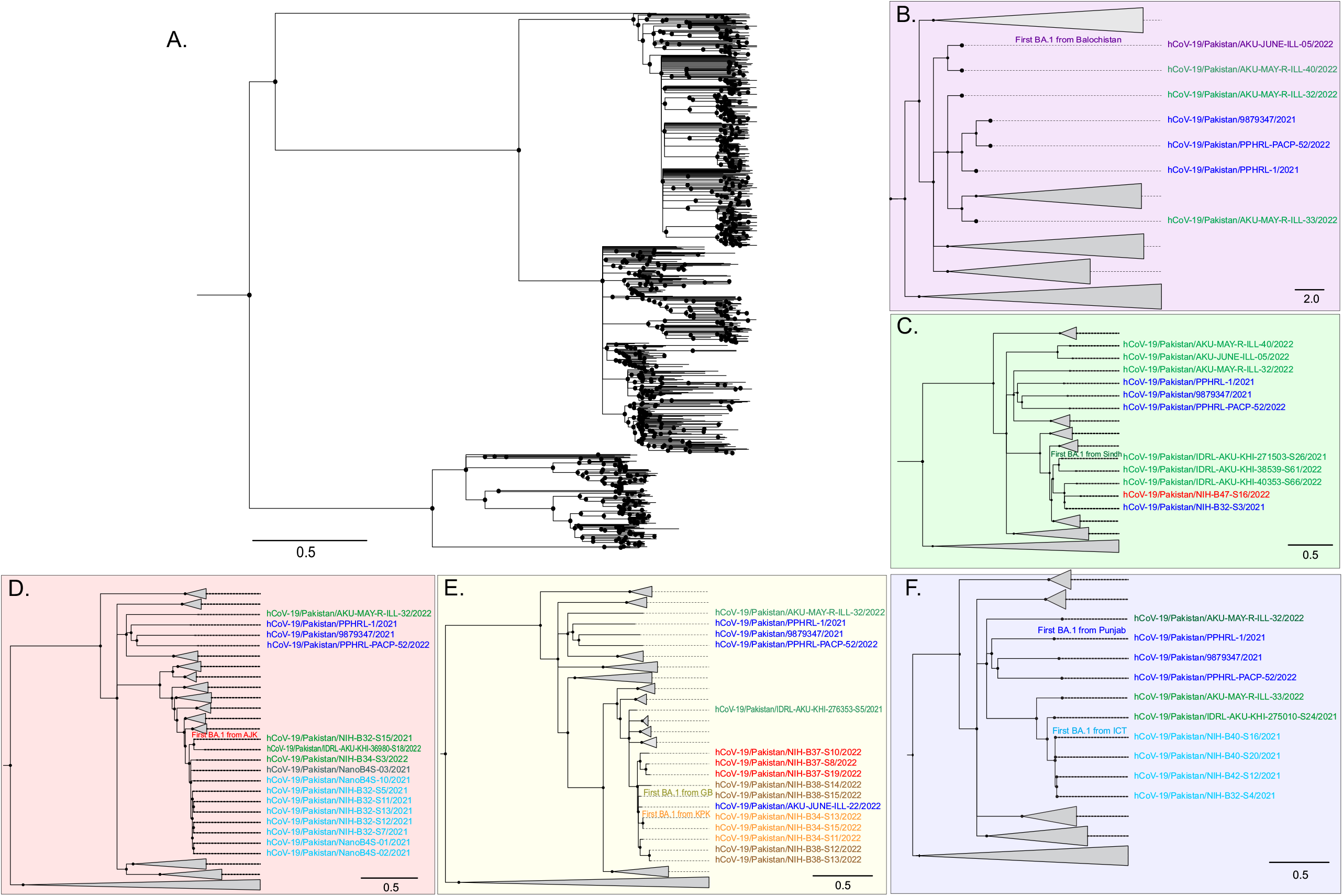
Evolutionary relatedness of the first reported Omicron variant from each region of Pakistan. A. Phylogenetic tree for 957 Omicron genomes and related strains sharing the most recent common ancestor (MRCA). The first BA.1 sequence from each region, B, Balochistan (purple); C, Sindh (green); D, AJK (light red); E, Gilgit-Baltistan and KPK (yellow); F, Punjab and ICT (light blue). Data are presented as Figtree *v*. 1.4.4 outputs using the maximum-likelihood tree generated using IQTREE *v*. 2.2.0. Grey triangles represent collapsed nodes of relatively distant sequences from the first Omicron sequences in each region. Scale bar presents nucleotide substitution/site.

## Discussion

Genomic surveillance data from Pakistan have been limited, especially for the earlier pandemic waves of 2020 and 2021. Our study provides insights into the phylogenetic relatedness of different Omicron variants which spread across Pakistan. Here, we associate the introduction of the Omicron subvariants with COVID-19 surges in the different regions across the country. This is the first study highlighting the genetic relatedness of Omicron subvariants in Pakistan through an analysis of the MRCAs. We identify Sindh as a hotspot for variant introductions into the country.

COVID-19 waves were found to display a pattern of sequential geographic transmission, occurring first in Sindh, followed by Punjab and other regions. Overall, the majority of COVID-19 cases were aged below 40 years. The predominant age group of cases was 19-40 years.

Deaths have previously been associated with older age groups [22]. During the period covered by this study, the CFR was found to be the highest in Balochistan (1.5%), 1.1% in KP and with all other regions reporting a CFR of less than 1%. Over the course of the COVID-19 pandemic, the highest mortality (3.5%) reported in Pakistan was from Peshawar, KP, in 2020. Local experts have suggested the lack of social distancing and a non-compliant response to standard operating procedures (SOPs) in the community to be the major reason behind this [23]. Other factors potentially contributing to the higher CFR could include a delayed presentation of COVID-19 symptoms combined with limited testing and reporting.

Balochistan reported the fewest cases, and the fewest genomic sequences were submitted from the region. Hence, due to the limited data available it is not possible to understand fully the trend of COVID-19 in Balochistan. One study showed COVID-19 positivity of 13% between March and December 2020, with the mean age of positive cases to be 36 ± 14 years, with 20% of the cases being female [24]. The lack of a provincial sequencing facility in Balochistan limits the available sequencing efforts. Another challenge regarding COVID-19 case information in Balochistan is the limited access to healthcare facilities for this large but sparsely populated province (6% of population with 43.6% of the land area of Pakistan).

Omicron variants comprised most of the sequences submitted to GISAID during the study period. BA.5 and BA.2 variants were the most prevalent overall amongst the genomes analyzed. We tried to determine if there was an association between the introduction of the first Omicron subvariant and the increase in number of cases across geographical regions of Pakistan. The first introduction was BA.1 on 8th December 2021 into Karachi, Sindh. BA.1 was reported later from Punjab, ICT and AJK. BA.1 reports from Balochistan and KP occurred after January 2022. Soon after identification of BA.1 strains, Pakistan experienced a surge of cases across the country, with particularly high numbers of cases and associated deaths in Karachi, Sindh. The identification of a larger than average case count in the Sindh region was associated with travelers from other destinations. Earlier, the introduction of the Alpha VoC was associated with international travelers to Karachi [25]. The high case count in Sindh could be also driven by the large population size; Karachi is a megacity with around 20 million inhabitants and, as the trading and financial hub of the country, receives more local and international travelers than other regions.

The first case from Sindh appeared to have transmitted locally as reported in previous studies [21]. The earliest BA.1 cases from ICT and Punjab share an ancestral node with Sindh, which does not necessarily mean that the variant spread through travel between these regions, but could also be attributed to an independent introduction from elsewhere.

Phylogenetic analysis showed that the first BA.1 strain from Balochistan was related to that from Sindh. The provinces have a shared border with frequent routine travel between them, allowing easy spread, however phylogeographic analysis would be needed to test this hypothesis.

The first BA.1 case from AJK (January 8, 2022) shared an ancestral node with a case from Sindh and Punjab. The earliest case from GB and KP had the same MRCA and shared an ancestral node with isolates from the Punjab. BA.1 cases in GB, KP, and AJK regions shared a same ancestral node, which might be due to the proximity of these three provinces in the northern region of Pakistan.

The first BA.2 variant was reported in January 2022 from Karachi, Sindh, and clustered closely with the isolate identified from a traveler from India. BA.2 variants were subsequently reported in ICT and then Punjab. BA.4 was first reported from AJK (May 2022) and BA.5 from Punjab (May 2022). The reporting of newly introduced variants was followed by local transmission. This was evident from reports in the same city/location in addition to those from other provinces.

It is likely that the rise in cases observed in Sindh (July 2022), Punjab (August 2022) and similar trends observed in other regions was due to the spread of BA.4 and BA.5 strains at this time. Notably, the increase in cases was first reported in Sindh. This is likely due to both its population size (mainly Karachi) and the extensive network of diagnostic laboratories in the region. The case numbers of the July–August 2022 surge were less than those of the January–February 2022 surge, possibly due to reduced testing rates within the population. Limited testing during the latter period could be attributed to reduced disease severity of COVID-19 from Omicron variants and increased vaccination coverage and thus less concern in the population about symptomatic infections.

There is a dearth of information regarding SARS-CoV-2 genomic surveillance from Asia, with studies providing insights into viral transmission from limited datasets[26]. This is the first study to conduct a detailed phylogenetic analysis of Omicron subvariants in Pakistan to understand their first introductions. Sindh had the highest number of reported COVID-19 cases (41.7% of total cases), while its population size is less than half of Punjab’s, suggesting that the rate of COVID-19 cases on a population basis was higher in Sindh than in Punjab.

Similarly, Balochistan had the lowest number of reported COVID-19 cases (0.9% of total cases), but its population size is also the smallest among all regions. However, the CFR in Balochistan was the highest (1.5%) among all regions, indicating that the pandemic’s impact was severe there. In a similar manner, KP has a population of 36 million but has much higher deaths as compared to Sindh (48 million) and Punjab (110 million), as reflected by the CFR (1.1%)

The availability of sequencing data varies widely between different regions in Pakistan. It is a limitation of this study that full interpretations of the impact of Omicron subvariants in the different provinces of Pakistan cannot be made due to limited genomic surveillance in some regions. For instance, despite having identified Sindh as the entry-point of viral strains, this could be skewed by the limited data from other provinces. Another consequence of the limited genomic surveillance is delayed sampling, testing, and reporting. We used the sample collection dates for phylodynamic analysis and are able to provide insights regarding strain variations across the study period. Additionally, the sequencing was not consistent over the entire study period, as more samples were sequenced in the wave between December 2021 and February 2022. This makes it difficult to analyze the data in the context of burden of disease with age and gender stratification. However, given that most of the COVID-19 cases reported from Pakistan are from those aged 40 years and below, it is not surprising that we found that there was a greater representation of Omicron variants in this age group. This is in keeping with the younger age of the population of Pakistan, with 65% of individuals aged below 30 years. Another limitation of the study is that only the first Omicron subvariant, BA.1, was studied in terms of phylogenetic relatedness. Overall, it is likely that BA.5 samples had a greater representation in this study selection as the increase in this VoC occurred during a period in 2022 when there had been an increase in sequencing capacity for SARS-CoV-2 genomics. In the context of Aga Khan University, this was due to a contribution of institutional (Aga Khan University), national (Higher Education Commission, Pakistan; World Health Organization, Pakistan) and international (Health Security Partners, USA; Fogarty International Center, NIH, USA; Bill and Melinda Gates Foundation) funding support for sequencing and bioinformatics initiatives.

In conclusion, correlation of SARS-CoV-2 genomic data with COVID-19 epidemiological data in Pakistan allowed us to describe the introduction of Omicron subvariants with different pandemic waves. Further, information regarding the relatedness of the lineages introduced in each province provides insights into the possible reasons for the waves observed in the provinces, which were led by Sindh and then followed in each case by ICT and Punjab. There is a need to establish more robust genomic surveillance networks to adequately represent the entire country. The importance of continuing genomic surveillance matched with analyzing epidemiological data is essential for successful management of a highly transmissible pathogen such as SARS-CoV-2.

## Data Availability

All data produced in the present work are contained in the manuscript. Further data can be asked from the corresponding author

## Acknowledgements

Research support was provided by the Aga Khan University, Pakistan; World Health Organization, Islamabad, Pakistan; Higher Education Commission, Pakistan; Health Security Partners, USA; and a Bill and Melinda Gates Foundation grant. Fogarty International Center, National Institutes of Health, USA provided training for genomic epidemiology. The opinions expressed in this article are those of the authors and do not reflect the view of the National Institutes of Health, the Department of Health and Human Services, or the United States government.

## Authors contributions

ARB, YAR, JA and ZH planned the study. ARB, AK, JA, PMT, BM, NST, WUK were involved in sequencing and bioinformatics analysis. YAR, AK, MY and ARB conducted the data analysis. ZH, RH, IN, WUK, ZR, DS and UBA got funding support for this work. ZR, NST and DS edited the paper. All authors approved the manuscript.

