## Supplementary figures and images for "Sequential viral introductions and spread of BA.1 drove the Omicron wave across Pakistani provinces"

### Supplementary Figure 1

**Supplementary Figure 1**

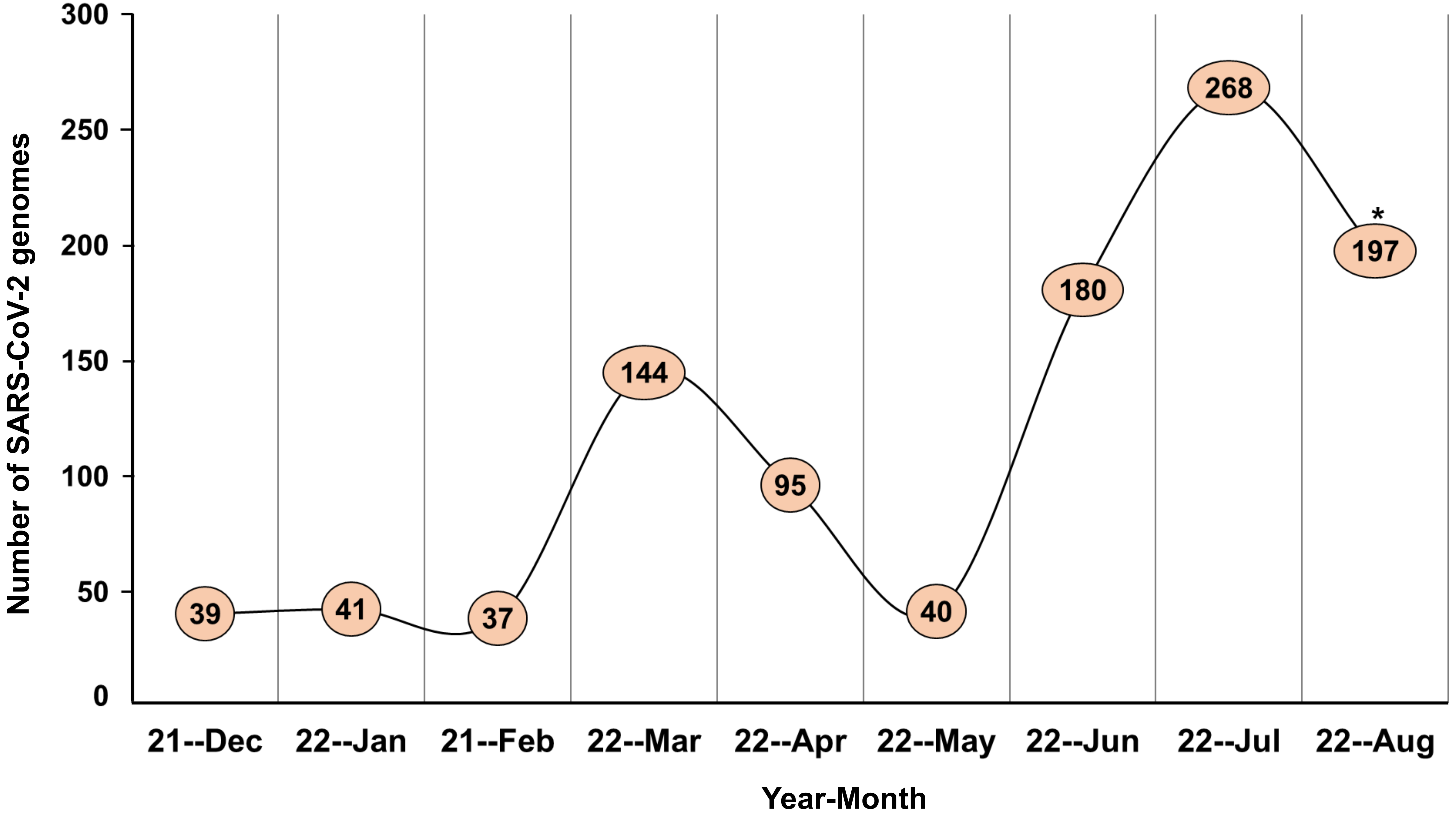

### Supplementary Figure 2

**Supplementary Figure 2**

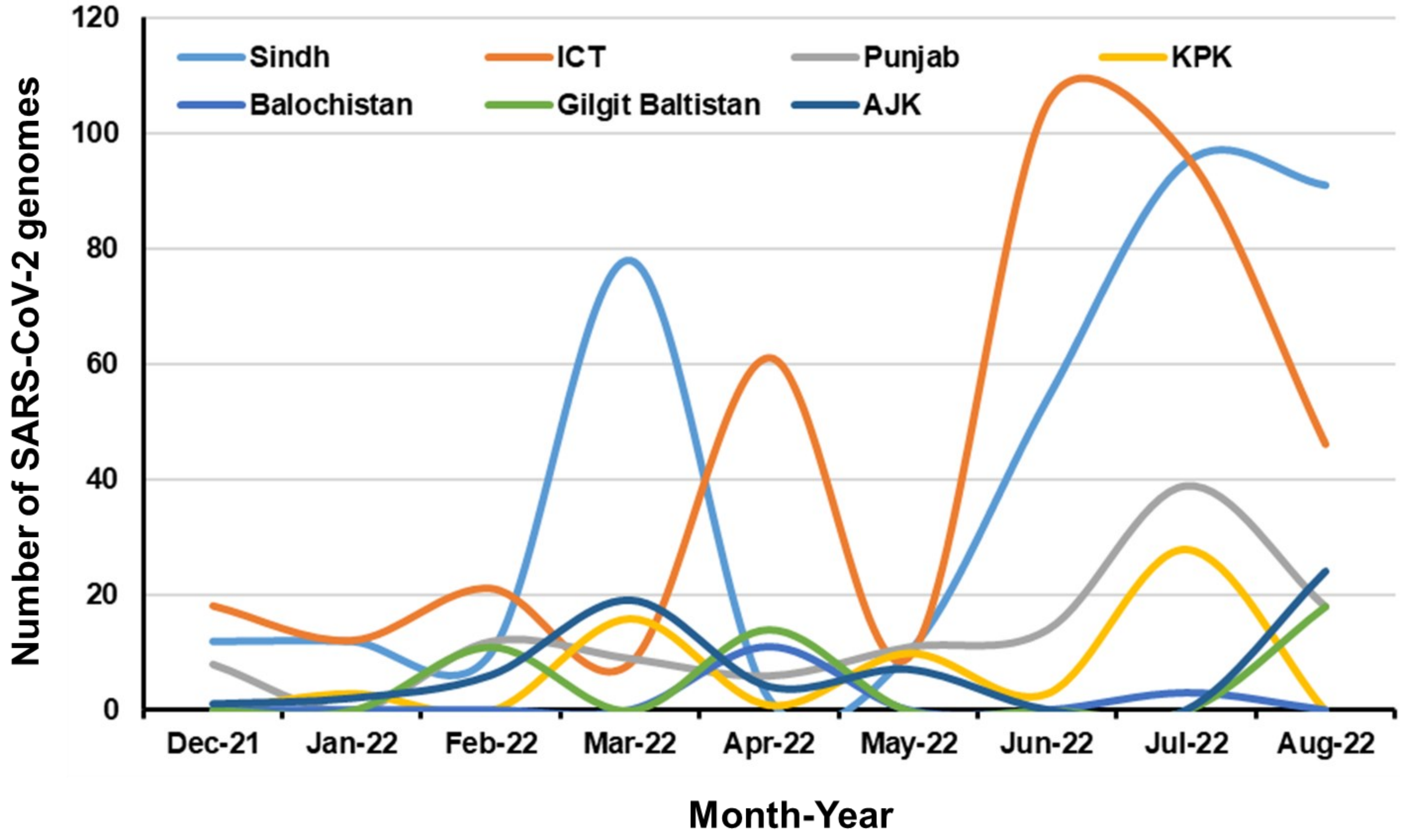
