## Supplementary Figure Legends for "Sequential viral introductions and spread of BA.1 drove the Omicron wave across Pakistani provinces"

**Supplementary Figure Legend**

**Supplementary Figure 1. SARS-CoV-2 genome submissions in GISAID from Pakistan.** Distribution of 1041 sequences submitted by Pakistan from December 1, 2021 till August 14, 2022. On x-axis are the date in year-month format and y-axis represents the number of SARS-CoV-2 genome submissions in GISAID. Orange circles shows the exact number of sequences. *partial data for August (till August 14, 2022).

**Supplementary Figure 2.** SARS-CoV-2 genome submissions in GISAID from each region of Pakistan across time. The graph shows all SARS-CoV2 sequences (n=1041) genomes submitted from each region of Pakistan between December 1, 2021 and August 14, 2022. Sindh (light blue), ICT (orange), Punjab (grey), KPK (yellow), Balochistan (blue), GB (green), AJK (dark blue). On x-axis date in month-year format is mentioned and y-axis shows number of SARS-CoV2 genomes submitted in GISAID.
